# Investigating the causal effects of childhood and adulthood adiposity on later life mental health outcome: a Mendelian randomisation study

**DOI:** 10.1101/2023.05.09.23289512

**Authors:** Sweta Pathak, Tom G Richardson, Eleanor Sanderson, Bjørn Olav Åsvold, Laxmi Bhatta, Ben Brumpton

**Affiliations:** K.G. Jebsen Center for Genetic Epidemiology, Department of Public Health and Nursing, Norwegian University of Science and Technology, Trondheim, Norway; MRC Integrative Epidemiology Unit, Population Health Sciences, Bristol Medical School, University of Bristol, United Kingdom; Department of Endocrinology, Clinic of Medicine, St. Olavs Hospital, Trondheim University Hospital, Trondheim 7030, Norway; HUNT Research Centre, Department of Public Health and Nursing, NTNU Norwegian University of Science and Technology, Levanger, Norway; Clinic of Medicine, St. Olavs Hospital, Trondheim University Hospital, Trondheim, Norway

**Author notes:** **Corresponding authors:** Sweta Pathak, Ben Brumpton, Department of Public Health and Nursing, Faculty of Medicine and Health Sciences, NTNU, Norwegian University of Science and Technology, P.O. Box 8905, MTFS, NO-7491, Trondheim, Norway. Equal contributions (Laxmi Bhatta, Ben Brumpton).

## Abstract

**Background:** Obesity particularly during childhood is considered a global public health crisis and has been linked with later life health consequences including mental health.However, there is lack of causal understanding if childhood adiposity has a direct effect on mental health or has an indirect effect after accounting for adulthood body size.

**Objective:** To investigate the total and direct effect of childhood adiposity on later life anxiety and depression.

**Method:** Two-sample Mendelian randomization (MR) was performed to estimate the total effect and direct effect (accounting for adulthood body size) of childhood body size on anxiety and depression. We used summary statistics from a genome-wide association study (GWAS) of UK Biobank (n=453,169) and large-scale consortia of anxiety (Million Veteran Program) and depression (Psychiatric Genomics Consortium) (n=175,163 and n=173,005, respectively).

**Result:** Univariable MR did not indicate genetically predicted effects of childhood body size with later life anxiety (beta=-0.05, 95% CI=-0.13, 0.02), and depression (OR=1.06, 95% CI=0.94, 1.20). However, using multivariable MR, we observed that the higher body size in childhood reduced the risk of later life anxiety (beta=-0.19, 95% CI=-0.29, -0.08) and depression (OR=0.83, 95% CI=0.71, 0.97). Both univariable and multivariable MR indicated that higher body size in adulthood increased the risk of later life anxiety and depression.

**Conclusion:** Our findings suggest that the higher body size in childhood has a protective effect on later life anxiety and depression, if obesity is not present into adulthood. Higher body size in adulthood was a risk factor for later life anxiety and depression.

## Background

Obesity is a global pandemic with a prevalence that has tripled since 1975 and it is a leading risk factor for adverse health outcomes including cardiometabolic diseases (World Health Organization, 2010). Obesity particularly during childhood is considered a global public health crisis and has increased 10-fold since 1975 (World Health Organization, 2010, United Nations International Children’s Emergency Fund, 2017). Childhood obesity has been linked to several health outcomes including cardiometabolic diseases and premature mortality (Baer *et al*., 2005, Bibbins-Domingo *et al*., 2007, Biro and Wien, 2010, Caird *et al*., 2011, Lindberg *et al*., 2020a, Lindberg *et al*., 2020c).

According to WHO in 2019, 1 in every 8 people were living with mental disorder among which anxiety (301 million) and depression (280 million) are most common (World Health Organization, 2022) and are frequently comorbid with one another (Jacobson and Newman, 2014, Kalin, 2020). Observational studies have reported an association between childhood adiposity and childhood anxiety and depression (Lindberg *et al*., 2020c, Patalay and Hardman, 2019, Quek *et al*., 2017). However, no study has investigated the effect of childhood obesity on later life anxiety. Two longitudinal studies have explored the association between childhood adiposity and later life depression, and observed that obese children are at higher risk of depression in adulthood compared to normal weight children (Tyrrell *et al*., 2019).

Simmonds et. al. observed that around 55% of children with obesity went on to be obese in adolescence, meaning nearly half were only obese during their childhood (Simmonds *et al*., 2015). The high proportion of subsequent obesity however means that it is difficult to disentangle the association of childhood and adulthood adiposity. Several reports suggest that the early years of life play a crucial role for health and wellbeing in later life (Center on the Developing Child at Harvard University, 2010, United Nations International Children’s Emergency Fund, 2017) (Human Early Learning and Commission on Social Determinants of, 2007). So, it is important to know how childhood obesity affect later life mental health while addressing the influence of adulthood obesity. Using genetic variants for childhood and adulthood adiposity and a novel multivariable Mendelian randomization (MR) method, may help to answer whether childhood obesity has direct effect on later life anxiety and depression or has a indirect effect after accounting for adulthood adiposity (Sanderson *et al*., 2022b).

MR studies use genetic variants as instrumental variable to test causal relationships between risk factors and outcomes and are typically considered to be more robust to counfounding and reverse causation often seen in traditional observational studies (Davies *et al*., 2018, Sanderson *et al*., 2022a). MR studies have suggested that adulthood adiposity may be a causal risk factor for later life anxiety and depression (Casanova *et al*., 2021, Hartwig *et al*., 2016a, He *et al*., 2022, Speed *et al*., 2019, van den Broek *et al*., 2018, Walter *et al*., 2015, Wray *et al*., 2018). However, no MR study to the date has investigated the causal effect of childhood adiposity on later life anxiety and depression. Hence, in this study we aim to estimate total and direct effect (i.e. after accounting for adulthood body size) of childhood adiposity on later life anxiety and depression.

## Methods

### Study design

We used a two-sample univariable and multivariable MR approach using summary-level data of genetic variants of childhood body size, adulthood body size, anxiety, and depression (Hartwig *et al*., 2016b) (Figure 1).

**Figure 1.**
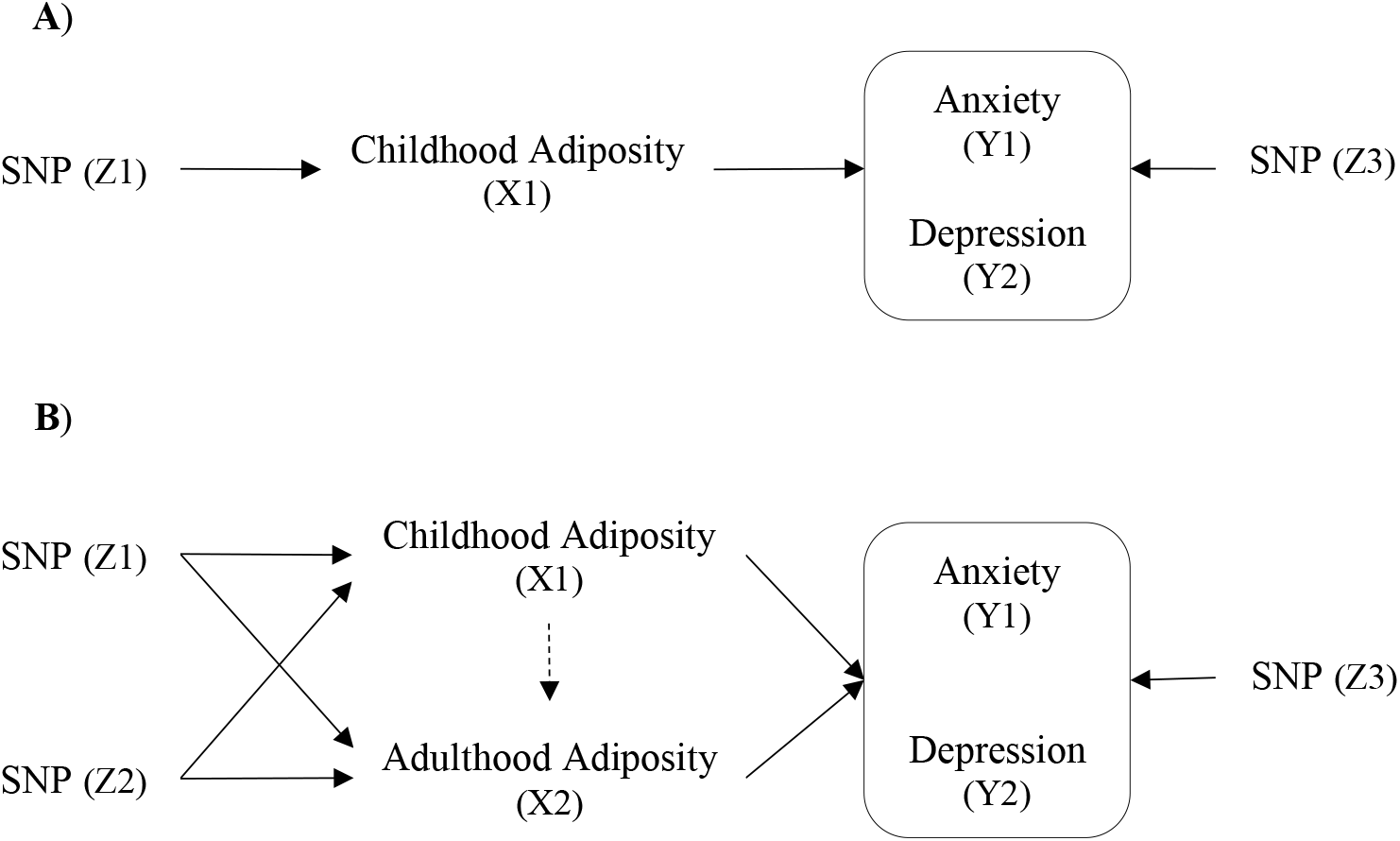
A directed acyclic graph of the two-sample MR framework. **A)** Univariable MR framework in the relationship of childhood adiposity on later life anxiety and depression. **B)** Multivariable MR framework where the solid (direct) and dotted (total) are direct and total pathways in the relationship of childhood adiposity through adulthood adiposity on later life anxiety and depression. Z1 - genetic variants associated with the exposure of interest, Z2 - genetic variants associated with the second exposure, Z3 - genetic variants associated with the outcome, X1 – First exposure, X2 – Second exposure, Y1 and Y2 – outcome.

**Figure 2.**
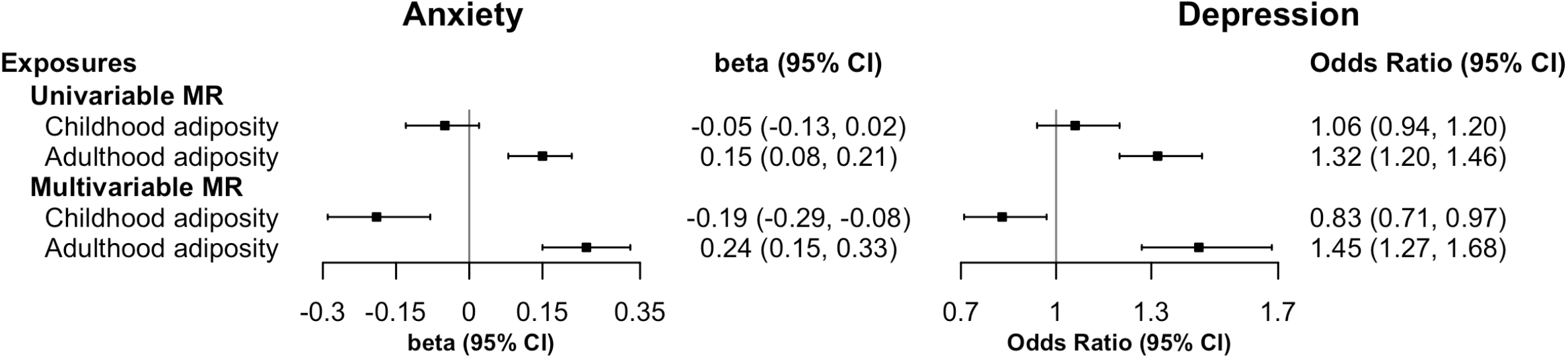
Forest plot illustrating the total and direct causal estimates of childhood adiposity and adulthood adiposity on anxiety and depression. The univariable MR represents the total causal estimate and multivariable MR represents direct causal estimates. The causal estimates are presented as beta for anxiety and odds ratio for depression with 95% CI.Abbreviations: MR, Mendelian randomization; CI, confidence interval

## Data resources

### Genetic instruments for childhood and adulthood body size

We retrieved SNP-exposure associations of childhood and adulthood body size from a GWAS performed in UK Biobank in 453,169 people with European ancestry adjusted for age, sex, genotyping chip, relatedness and population stratification (Richardson *et al*., 2020). In this study, 313 and 580 independent genetic variants were identified as genetic instrument for childhood and adult body size, respectively (Richardson *et al*., 2020, Zhao *et al*., 2022) using the clumping criteria of r^2^ <0.001 and p-value <5×10^−08^. Adult BMI was measured when people were between aged 40 and 69 years and at the same time people were asked to describe their childhood body size when they were 10 years as thinner, plumper, and about average (Richardson *et al*., 2020). Later to make both measures comparable, adult body size was also classified as thinner, plumper, and about average by categorising individuals into those groups in the same proportions as was observed in the childhood data. Validation and simulation analysis was carried out where only people with early life and later life measures were included to account for the limitation of using recalled childhood body size (Richardson *et al*., 2020). The genetic instrument of childhood body size was distinct from the genetic instrument of adulthood body size with a genetic correlation (rg) of 0.61 (Richardson *et al*., 2020). In a validation study, by using measured BMI data from independent studies in the Avon Longitudinal Study of Parents and Children (ALSPAC) (Richardson *et al*., 2020), the Young Finns study (Richardson *et al*., 2021) and the Trøndelag Health (HUNT) study (Brandkvist *et al*., 2021), the genetic instrument for childhood body size from UK Biobank was found to be strong predictor of childhood body mass index (BMI) and less predictive of adulthood BMI. The findings were validated by calculating the risk predictive ability of genetic score of childhood and adulthood genetic instruments.

### Anxiety and depression

The summary statistics for the SNP-outcome associations (anxiety and depression) were retrived from GWAS performed in large studies based on European ancestry. The GWAS of anxiety was performed in Million Veteran Program (MVP) among 175,163 people with mean age 66.58 years (Levey *et al*., 2020). Anxiety was measured using two questionnaires (Supplementary Table S7), later responses were summed and then scored on a continuous scale from 0 to 6 (based on score on the Generalized Anxiety Disorder 2-item scale [GAD-2] (Kroenke *et al*., 2010)). The GWAS analysis of anxiety were adjusted for age, sex, and the first 10 within ancestry principle component. Details of the GWAS on anxiety can be found elsewhere (Levey *et al*., 2020).

The GWAS meta-analysis of depression was performed in 173,005 people (59851 cases and 113154 control, European ancestry) from Psychiatric Genomics Consortium (PGC) including 29 cohorts and 5 additional independent cohort (Wray *et al*., 2018). Comparability of these cohorts was examined by genetic correlation of common variants between them. Major depression cases were assessed using traditional method [Diagnostic and Statistical Manual of Mental Disorders (DSM-IV) or International Classification of Disease (ICD-9, ICD-10)] and from treatment registers, which was measured on dichotomous scale. Details of the GWAS on depression can be found elsewhere (Wray *et al*., 2018).

### Statistical Analysis

We used univariable MR to estimate the total effect of genetically predicted childhood and adult body size on anxiety and depression. We used multivariable MR to estimate the direct effect of genetically predicted childhood body size conditioning on adult body size and vice-versa on anxiety and depression. The ‘MendelianRandomization’ package in R statistical software was used. In both univariable and multivariable analyses, effects were estimated using the inverse variance weighted (IVW) method. IVW estimates are obtained by combining each individual genetic variant associations, such individual association estimate are obtained by dividing its gene-outcome association by its gene-exposure association, where the weight of each ratio is the inverse of the variance (MR Dictionary). If all the genetic variants used in MR are valid, the IVW estimate is most powerful (Burgess *et al*., 2013).

The risk alleles of childhood body size were harmonized with the risk alleles of adulthood body size and outcome (anxiety and depression). Finally a harmonized data set that contain the same allele pairs were analyzed. We calculated conditional F statistic to test the strength of genetic variants of childhood and adulthood body size (Sanderson *et al*., 2021). Valid instruments can provide independent and unbiased causal estimates of exposure and outcome (Cho *et al*., 2020). The conditional F-statistics of childhood body size conditioned on adulthood body size was >10 suggesting that potential bias due to weak instruments was unlikely (Supplementary Table S2).

As a sensitivity analysis to evaluate the validity of IVW results in the univariable and multivariable MR analysis, we calculated MR Egger intercepts and estimates, and additionaly for the univariable MR analysis, weighted mode and weighted median was performed to explore potential horizontal pleiotropy (that is when genetic variants for the exposure of interest has an effect on outcome through a phenotype other than the exposure).

We presented the estimates in forestplots using the ‘forestplot’ package in R.

## Results

In univariable MR analysis, there was no evidence of a total effect from genetically predicted childhood adiposity on later life anxiety (beta= -0.05 per change in body size, 95% CI =-0.13, 0.02) and depression (OR= 1.06 per change in body size, 95% CI = 0.94, 1.20). Whereas, for genetically predicted adulthood adiposity there was strong evidence of a positive effect on anxiety (beta= 0.15 per change in body size, 95% CI = 0.08, 0.21) and depression (OR= 1.32 per change in body size, 95% CI = 1.20, 1.46).

In multivariable MR analysis, there was evidence of a direct effect of childhood adiposity (accounting for adulthood adiposity) with a strong protective effect on anxiety (beta= -0.19 per change in body size, 95% CI = -0.29, -0.08) and depression (OR= 0.83 per change in body size, 95% CI = 0.71, 0.97). Moreover, the direct effect of adulthood body size (accounting for childhood body size) was consistent with univariable MR with a strong increase in anxiety (beta= 0.24 per change in body size, 95% CI = 0.15, 0.33) and depression (OR= 1.45 per change in body size, 95% CI = 1.27, 1.68).

Statisical evidence of heterogeneity was observed across all analyses (supplementary Table S3). In univariable MR, MR egger intercept provide no evidence of horizontal pleiotropy. However in a sensitivity analysis, MR egger estimates for adulthood adiposity slightly vary from IVW estimates. In multivariable MR, MR egger intercept provide no evidence of horizontal pleiotropy and MR egger estimates were similar to IVW estimates (supplementary table S4 and S5).

## Discussion

In this MR study, we examined the influence of early life body size (age 10 year) on later life anxiety and depression. Our multivariable MR estimates suggest that genetically predicted larger body size at childhood has a direct effect reducing the risk of later life anxiety and depression, provided that people do not continue to be obese as adults. However, higher body size in adulthood was a risk factor for later life anxiety and depression.

To our knowledge, no study has investigated the association of childhood adiposity on later life anxiety. However, Lindberg et. al. observed a positive association between childhood adiposity and childhood anxiety in a nationalwide Swedish study (Lindberg *et al*., 2020b). In contrast, a systematic review and meta-analysis by Moradi et al. observed no association between childhood adiposity and childhood anxiety (Moradi *et al*., 2022). Several other studies have suggested inconsistent associations between childhood obesity and childhood anxiety, but no studies have reported a protective association (Godina-Flores *et al*., 2022, Hughes *et al*., 2022, Mazurak *et al*., 2021). Therefore, further validation of our findings in an independent study is warrented.

Only two observational studies have examined the association of childhood adiposity on later life depression. Tyrrell et. al. (n=287,503) studied the UK biobank population and observed that the thinner and plumper child has increased risk of depression compared to child with average body size (Tyrrell *et al*., 2019). Gibson-Smith et. al. (n=889) observed increased risk of major depressive disorder (MDD) among obese child compared to child with normal body size; however, did not observed any association between childhood obesity and later life depressive symtoms (Gibson-Smith *et al*., 2020). In both of studies by Tyrrell et. al. and Gibson-Smith et. al., the effect of childhood adiposity remained the same even after adjusting for adulthood BMI (Gibson-Smith *et al*., 2020, Tyrrell *et al*., 2019). In contrast to these findings from observational studies, our study suggests a protective effect of childhood body size on later life depression after adjusting for adulthood body size using a multivariable MR approach. A possible reason for the differences in results might be due to the differences in study design, where we used a MR approach, which may have avoided residual confounding which commonly affects observational studies (Davies *et al*., 2018).

Besides our study, no other multivariable MR studies have examined the role of childhood adiposity adjusting for adulthood adiposity on risk of later life anxiety and depression.

However, several multivariable MR studies have investigated the effect of genetically predicted childhood body size adjusting for adulthood body size on different later life health outcomes. Larger body size during childhood has been observed to increase the risk of health outcomes such as coronary artery disease, type 1 diabetes, colorectal cancer, cardiovascular disease, and heart structure (O’Nunain *et al*., 2022, Papadimitriou *et al*., 2023, Richardson *et al*., 2022, Richardson *et al*., 2020). However, protective effects of genetically predicted higher body size during childhood have been observed for fracture and breast cancer in later life (Power *et al*., 2021, Richardson *et al*., 2020). A similar protective effect for mental health (anxiety and depression) was observed by our study for childhood adiposity, while an increased risk for poor mental health was observed for adulthood body size. These findings suggest that larger body size in childhood is not contributing to later life poor mental health, but it is large body size in adulthood that is the risk factor. Additionally, the findings highlight the public health importance of a life course approach in our understanding of adverse health outcomes, and how the quantification of risk/benefits of adiposity at different stages of life could guide targeted life course intervention and improve diseases prevention. However having large body size during adulthood increases the risk of poor mental health.

A strength of our study is that the two-sample MR framework allowed us to use genetic instruments from GWAS with the largest available sample sizes, thus improving statistical power. In addition, in the two-sample MR approach, we used mostly non-overlapping data sets for the exposure and outcome which helps to reduce potential bias from overfitting (Burgess *et al*., 2016). This study used the novel multivariable MR approach to quantify the life course effect of childhood body size on later life mental health, which has used genetic variants for body size at two specific time periods over the lifecourse (Burgess and Thompson, 2015, Sanderson, 2021, Sanderson *et al*., 2019). This novel approach allowed us to avoid pleiotropy cancelling out opposing effects from the childhood and adulthood SNPs and therefore capture the effect of childhood adiposity and mental health, when it was not observed in univariable analysis.

However, the results should be interpreted in light of certain limitations. The UK biobank contribute 17% of the total sample size in the depression GWAS used for our analysis, which is a small contribution to the overall GWAS estimates but might introduce biases towards the observational estimates in our two-sample MR analysis. The childhood body size is self-reported body size, where adult participants (average age 56.5) recalled their childhood body size when they were 10 year old. This might have introduced some recall bias. Although simulations showed that this was unlikely (Tom *et al*., 2019). Additionally, as describe earlier, three validation studies in different cohorts were conducted confirming the genetic instrument for perceived childhood body size (Brandkvist *et al*., 2021, Richardson *et al*., 2021, Richardson *et al*., 2020). We cannot however rule out that the underlying pathophysiology of childhood and adulthood adiposity is the same and therefore may reflect different pathways to disease which may complicate our interpretation. Our univariable and multivariable MR results are also vulnerable to confounding due to population stratification, dynastic effects and assortative mating (Brumpton *et al*., 2020). A further limitation of our study is the lack of ancestral diversity which limits the generalizability of results. Therefore, future research is warranted with broader range of different ancestries. Finally, compared to the general population, UK biobank participants are more likely to be older, female and live in less socioeconomically deprived areas which might have introduced selection bias in our study (Fry *et al*., 2017).

## Conclusion

Using multivariable Mendelian randomization, our findings provide novel evidence that higher body size during childhood (accounting for adulthood body size) has a protective effect on later life anxiety and depression, provided that normal body size during adulthood is maintained. In contrast, having larger body size during adulthood (accounting for childhood body size) may increase the risk of anxiety and depression. Our study highlights the need for future studies to consider the impact of adiposity at different time points in life on later life anxiety and depression.

## Data Availability

1) Summary statistic for childhood and adulthood adiposity are available online.
2) Summary statistic for depression are available online.
3) Summary statistic for anxiety are available uopn request to the author through db GaP.

https://rmdopen.bmj.com/content/8/2/e002321

https://pubmed.ncbi.nlm.nih.gov/31906708/

https://www.nature.com/articles/s41588-018-0090-3

## Acknowledgements

We thank the participants of the UK Biobank study and the genome-wide association study consortia who made their summary statistics publicly available for this study.

## Author contributions

S. P., L. B. and B. B. conceived and design the study. S. P. analysed the data and wrote first draft of the manuscript. All authors took part in the interpretation and revision of the manuscript. S. P. and B. B. are accountable for the accuracy and integrity of all parts of the works.

## Ethical standards

We used publicly available summary statistics from published studies. Ethical approval and participant consent for each study is detailed in the respective publications (Levey *et al*., 2020, Wray *et al*., 2018, Zhao *et al*., 2022).

