## Supplementary material for "Investigating the causal effects of childhood and adulthood adiposity on later life mental health outcome: a Mendelian randomisation study": Table S1

[#] **Equal contributions**

* **Corresponding authors**:

**Table S1.** Causal estimates of childhood and adulthood adiposity on anxiety and depression.

| Exposures | Outcomes | Univariable MR | | | | Multivariable MR | | | |
|---|---|---|---|---|---|---|---|---|---|
| | | **SNPs** | **Beta/OR*** | **95% CI** | **P-value** | **SNPs** | **Beta/OR*** | **95% CI** | **P-value** |
| Childhood adiposity | Anxiety | 277 | -0.05 | -0.13 to 0.02 | 0.171 | 652 | -0.19 | -0.29 to -0.08 | $3.86\times10^{-04}$ |
| | Depression | 260 | 1.06 | 0.94 to 1.20 | 0.345 | 638 | 0.83 | 0.71 to 0.97 | 0.019 |
| Adulthood adiposity | Anxiety | 510 | 0.15 | 0.08 to 0.21 | $1.11\times10^{-05}$ | 652 | 0.24 | 0.15 to 0.33 | $1.09\times10^{-07}$ |
| | Depression | 496 | 1.32 | 1.20 to 1.46 | $3.31\times10^{-08}$ | 638 | 1.45 | 1.27 to 1.68 | $1.33\times10^{-07}$ |

Table illustrating the total (univariable MR) and direct (multivariable MR) causal estimates of childhood and adulthood adiposity on later life anxiety and depression.

*The causal estimates are presented as beta for anxiety and odds ratio for depression with 95% CI and P-value. Number of SNPs (genetic variant) involve in analysis to investigate causal effect of exposure on outcome also presented.

Abbreviations: MR, Mendelian randomization; CI, confidence interval; SNP, Single nucleotide polymorphism; OR, Odds ratio

**Table S2**. Conditional F statistics in univariable and multivariable MR

| Exposure | Outcome | Univariable MR | Multivariable MR |
|---|---|---|---|
| Childhood adiposity | Anxiety | 65.35 | 13.34 |

| | | | |
|---|---|---|---|
| | Depression | 66.03 | 13.47 |
| Adulthood adiposity | Anxiety | 52.14 | 15.74 |
| | Depression | 52.43 | 15.60 |

**Table S3**. Heterogeneity test of univariable and multivariable MR

| **Exposure** | **Outcome** | **Univariable MR** | **Multivariable MR** |
|---|---|---|---|
| | | **Q (P-value)** | **Q (P-value)** |
| Childhood adiposity | Anxiety | 443 ($7.34\times10^{-10}$) | 1076 ($1.46\times10^{-23}$) |
| | Depression | 527 ($1.76\times10^{-20}$) | 1134 ($1.40\times10^{-30}$) |
| Adulthood adiposity | Anxiety | 885 ($1.05\times10^{-22}$) | 1076 ($1.46\times10^{-23}$) |
| | Depression | 874 ($2.18\times10^{-23}$) | 1134 ($1.40\times10^{-30}$) |

**Table S4.** Univariable and multivariable MR Egger estimates of childhood and adulthood adiposity on anxiety and depression.

| **Exposures** | **Outcomes** | **Univariable MR** | | | | **Multivariable MR** | | | |
|---|---|---|---|---|---|---|---|---|---|
| | | **SNPs** | **Beta/OR** | **95% CI** | **P-value** | **SNPs** | **Beta/OR** | **95% CI** | **P-value** |
| Childhood adiposity | Anxiety | 277 | -0.03 | -0.19 to 0.13 | 0.702 | 652 | -0.17 | -0.29 to -0.06 | 0.004 |
| | Depression | 260 | 1.18 | 0.91 to 1.55 | 0.216 | 638 | 0.85 | 0.71 to 1.02 | 0.085 |

| | | | | | | | | | |
|---|---|---|---|---|---|---|---|---|---|
| Adulthood adiposity | Anxiety | 510 | 0.02 | -0.17 to 0.20 | 0.854 | 652 | 0.26 | 0.16 to 0.35 | $3.40\times10^{-07}$ |
| | Depression | 496 | 1.11 | 0.83 to 1.47 | 0.489 | 638 | 1.49 | 1.28 to 1.74 | $2.93\times10^{-07}$ |

**Table S5.** Univariable and multivariable MR Egger intercepts of childhood and adulthood adiposity on anxiety and depression.

| **Exposures** | **Outcomes** | **Univariable MR** | | | **Multivariable MR** | | |
|---|---|---|---|---|---|---|---|
| | | **SNPs** | **intercept** | **P-value** | **SNPs** | **intercept** | **P-value** |
| Childhood adiposity | Anxiety | 277 | -0.0002 | 0.784 | 652 | 0.000 | 0.583 |
| | Depression | 260 | -0.002 | 0.367 | 638 | -0.001 | 0.432 |
| Adulthood adiposity | Anxiety | 510 | 0.002 | 0.142 | 652 | 0.001 | 0.583 |
| | Depression | 496 | 0.002 | 0.185 | 638 | -0.001 | 0.432 |

Table S6. Univariable weighted mode and weighted median of childhood and adulthood adiposity on anxiety and depression

| **Exposures** | **Outcomes** | **Univariable weight median** | **Univariable weighted mode** |
|---|---|---|---|

| | | SNPs | Beta/OR | 95% CI | P-value | SNPs | Beta/OR | 95% CI | P-value |
|---|---|---|---|---|---|---|---|---|---|
| Childhood adiposity | Anxiety | 277 | 0.006 | -0.10 to 0.11 | 0.900 | 277 | 0.04 | -0.11 to 0.21 | 0.589 |
| | Depression | 260 | 1.20 | 0.02 to 0.36 | 0.032 | 260 | 1.28 | 0.05 to 0.44 | 0.014 |
| Adulthood adiposity | Anxiety | 510 | 0.12 | 0.02 to 0.22 | 0.016 | 510 | 0.16 | -0.02 to 0.34 | 0.08 |
| | Depression | 496 | 1.43 | 0.23 to 0.50 | $5.34\times10^{-07}$ | 496 | 1.58 | 0.16 to 0.75 | 0.003 |

Table S7. GAD-2 Phenotype

| **GAD-2** | | | | |
|---|---|---|---|---|
| Over the last 2 weeks, how often have you been bothered by the following problems ? (Use "√" to indicate your answer) | Not at all | Several days | More than half the days | Nearly every day |
| Feeling nervous, anxious, or on edge | 0 | 1 | 2 | 3 |
| Not being able to stop or control worrying | 0 | 1 | 2 | 3 |
